# Analysis of corneal real astigmatism changes and high order aberration after lower eyelid epiblepharon repair surgery

**DOI:** 10.1101/19006080

**Authors:** Dong Cheol Lee

## Abstract

**Objective:** To investigate changes in corneal low and high order aberrations (LOA and HOA), which cause visual disturbances after lower eyelid epiblepharon repair surgery.

**Methods and analysis:** This was a retrospective, cross-sectional study, which included 108 eyes from 54 patients. Wavefront analyses for calibrated LOAs and HOAs (root mean square, coma, three-piece aberrations [Trefoil], secondary astigmatisms, and spherical aberrations [SA]) were performed via a Galilei G4 Dual Scheimpflug Analyzer preoperatively, at the first and second follow-ups (f/u), and at G1, G2, and G3 (<45 days, 45–75 days, and >75 days post-surgery). Several risk factors (age, sex, body mass index, and corneal keratitis presence) were assessed.

**Results:** In LOAs, flat keratometer (K) and axis values decreased significantly from baseline at the first f/u. At the second f/u, mean K and axis decreased. In HOAs, coma and trefoil increased from baseline at the first f/u and normalized by the second f/u. SA decreased at the second f/u and in G3. The various risk factors did not affect postoperative outcomes, axis, and secondary astigmatisms. After correcting for risk factors, at the first f/u, cylinder, coma, trefoil, and SA increased significantly from the baseline, while axis and flat K decreased. At the second f/u, cylinder increased, while axis and mean K decreased significantly from the baseline.

**Conclusion:** Epiblepharon repair surgery may impact axis changes. Flat K, coma, and trefoil may be affected by mechanical force changes immediately following surgery. Mean K and SA may change with cornea state changes during healing.

## INTRODUCTION

Epiblepharon is a congenital eyelid anomaly that more often affects Asian children than children of other demographics.[1] The prevalence of epiblepharon in the Japanese population was reported to be 9.9% in children aged 3 months to 18 years.[2] In addition, epiblepharon comprised 9.5% of the oculoplastic surgery clinical cases in a tertiary care hospital setting in Singapore.[3] Epiblepharon is defined as a horizontal fold in the hypertrophied pretarsal orbicularis muscle and redundant skin near the eyelid margin, which causes the eyelashes to project inwards towards the cornea while the eyelid and tarsal plate positions remain normal. Epiblepharon is differentiated from congenital entropion, which is defined as an inward rotation of the tarsus and the lid margin.[4, 5]

Patients with epiblepharon experience photophobia, foreign body sensation, irritating tearing, and visual disturbances caused by corneal problems.[6] In addition to these symptoms, astigmatisms are most frequently observed in patients with epiblepharon as compared with the normal population. For instance, Preechawai et al. reported a high prevalence of astigmatism in patients with epiblepharon (52.2% had a 1 D or worse astigmatism).[7] It is thought that the corneal curvature changes in epiblepharon are due to the mechanical force created by squeezing the eyelids in response to corneal erosion and abnormal eyelid tension from the hypertrophied pretarsal orbicularis muscle and redundant skin folds. Several studies have reported a greater prevalence of astigmatism in epiblepharon patients and changes in astigmatism after reparative surgery.[7-10]

Modern vision research has benefited from wavefront technology, which makes it possible to measure low- and high-order aberrations, including posterior corneal astigmatisms, with total cornea curvature via the Galilei G4 Dual Scheimpflug Analyzer (Ziemer, Port, Switzerland). Ray tracing using Snell’s law and pachymetry data with a reference plane in the posterior corneal surface are used.[11] Furthermore, higher order aberrations (HOAs) from all refractive errors, such as those that occur with astigmatisms, cannot be corrected by ordinary glasses or contact lenses. Recently, HOAs have been represented mathematically by calculating a Zernike coefficient.[12-14] Moreover, corneal transplantation, intraocular lens (IOL) implantation, pterygium surgery, and scleral buckling result in HOA changes.[15-17] In HOAs, ‘spherical aberration’ changes are known to result in decreased contrast sensitivity, halos, and glare, and ‘coma’ changes are associated with tilt and double vision.[18, 19] These HOAs result in changes that may lead patients to experience a qualitative deterioration in sight.[18-20]

Given that most epiblepharon patients are children, understanding the visual impairments caused by surgery is especially critical. The risk for astigmatism and HOAs due to surgery should be conveyed to patients, and eyeglass prescriptions should be considered. To our knowledge, although changes in anterior lower order aberrations (LOAs) (sphere and cylinder) after surgery have been investigated, no studies on the changes in calibrated anterior and posterior LOAs together with HOAs, including their risk factors, have been conducted. The purpose of the present study was to evaluate the preoperative and postoperative calibration of LOA (anterior and posterior astigmatisms) and HOA changes in the postoperative period, while adjusting for potential risk factors, such as age, sex, body mass index (BMI), and corneal keratitis status.

## METHODS

Approval for the retrospective review (IRB file No.: DSMC 2019-02-005) of our patients’ medical data was obtained from the Keimyoung University Dongsan Hospital Institutional Review Board (IRB), Daegu, Korea. All data and research collection procedures followed the tenets of the Declaration of Helsinki.

The medical and surgical records of 78 patients (156 eyes) diagnosed with epiblepharon who underwent epiblepharon repair surgery at Keimyoung University Dongsan Hospital from January 2016 to January 2019 were retrospectively reviewed. Due to the retrospective nature of this study, the IRB waived the requirement for patient consent. All surgeries were performed by a single surgeon (DCL), who was also involved in the clinical assessment of the patients. Surgical indications included severe corneal erosions and chronic irritating symptoms such as epiphora, frequent eye blinking and/or rubbing, and photophobia. Patients with post-surgery follow-ups of as long as 3 months were included. Patients with congenital entropion, distichiasis, trichiasis, or a history of ocular and eyelid surgery were excluded. A total of 48 eyes of 24 patients were excluded from this study.

Preoperatively, all subjects underwent ophthalmic examinations, including best-corrected visual acuity, cycloplegic refraction, slit-lamp examinations, and indirect fundus examinations. In addition, wavefront analyses for calibrated LOAs (anterior and posterior astigmatisms including keratometry were calibrated in ray-traced mode) and HOAs (root mean square [RMS], coma, three-piece aberrations [Trefoil], secondary astigmatisms [2^nd^ Astig], and spherical aberrations [SA]) were measured via a Galilei G4 Dual Scheimpflug Analyzer. These analyses were performed by a single technician preoperatively, and at the first and second f/u in group 1 (less than 45 days post-surgery), group 2 (from 45 to 75 days post-surgery), and group 3 (more than 75 days post-surgery).

### Surgical technique

All surgeries were performed under general anesthesia due to patient age. A mixture of lidocaine 2% and epinephrine (1:100,000) was injected subcutaneously into the lower eyelid. Excess lower eyelid skin was grasped with forceps to cause mild ectropion, and the lid was marked with a pen. The upper skin incision line was approximately 2 mm below the eyelash line, with a delineated ellipse more lateral than proximal to the punctum. The width of the ellipse was extended laterally near the lateral canthus to achieve a good contour.

The infra-eyelash skin incision and excision of the pretarsal orbicularis muscle was followed by suturing of the subcutaneous tissue of the upper skin-muscle flap to the exposed tarsal plate with three interrupted 6.0 monosyn sutures (medial, center, and lateral) to achieve a good eyelash contour. Moreover, a Bovie monopolar needle cautery instrument (Colorado Biomedical Inc, Evergreen, CO) was used for thermal cauterization of the septum to create adhesions between the preseptal orbicularis oculi muscle and the septum. These adhesions minimized vertical overriding of the orbicularis oculi muscle at the lower eyelid. If there were dog-ears at the lateral ends of the incision, the surgeon removed them with a triangular skin excision. If there were small amounts of pretarsal orbicularis oculi muscle and redundant skin overlying the lower lid margin, the tissue was also removed. At the end of surgery, the skin was then closed with a continuous 6-0 fast-absorbing plain gut suture. Ocuflox antibiotic ointment was used on the skin wound for 4 weeks postoperatively and was then tapered weekly.

### Statistical Methods

R language version 3.3.3 (R Foundation for Statistical Computing, Vienna, Austria) and T&F program version 2.9 (YooJin BioSoft, Korea) were used for all statistical analyses. Normally distributed variables are expressed as means ± standard deviations, and non-normally distributed variables are expressed as medians (interquartile range). For categorical variables, data are expressed as sample numbers and percentages (N [%]).

Paired sample t-tests were used to test for differences in outcomes from pre- to post-operation. When variables were not normally distributed, Wilcoxon signed rank tests were performed. Kolmogorov-Smirnov normality tests and Shapiro-Wilk normality tests were used to check the normality of all continuous variables.

Univariable linear regression analyses were performed to analyze the independent effects of risk factors such as age, sex, BMI, and cornea keratitis presence on continuous outcomes after surgery. For multivariable analysis, outcomes measured before and after surgery and risk factors such as age, sex, BMI, and cornea keratitis presence were used as paired dependent variables and independent variables, respectively, in a linear mixed-effect model. Time and all risk factors were used as fixed effect covariates with random intercepts across subjects. Statistical significance was reached when two-tailed *p*-values were less than 0.05.

## RESULTS

A total of 108 eyes of 54 patients were included in the study. Baseline patient characteristics, as well as the average f/u times, are shown in Table 1.

**Table 1.**
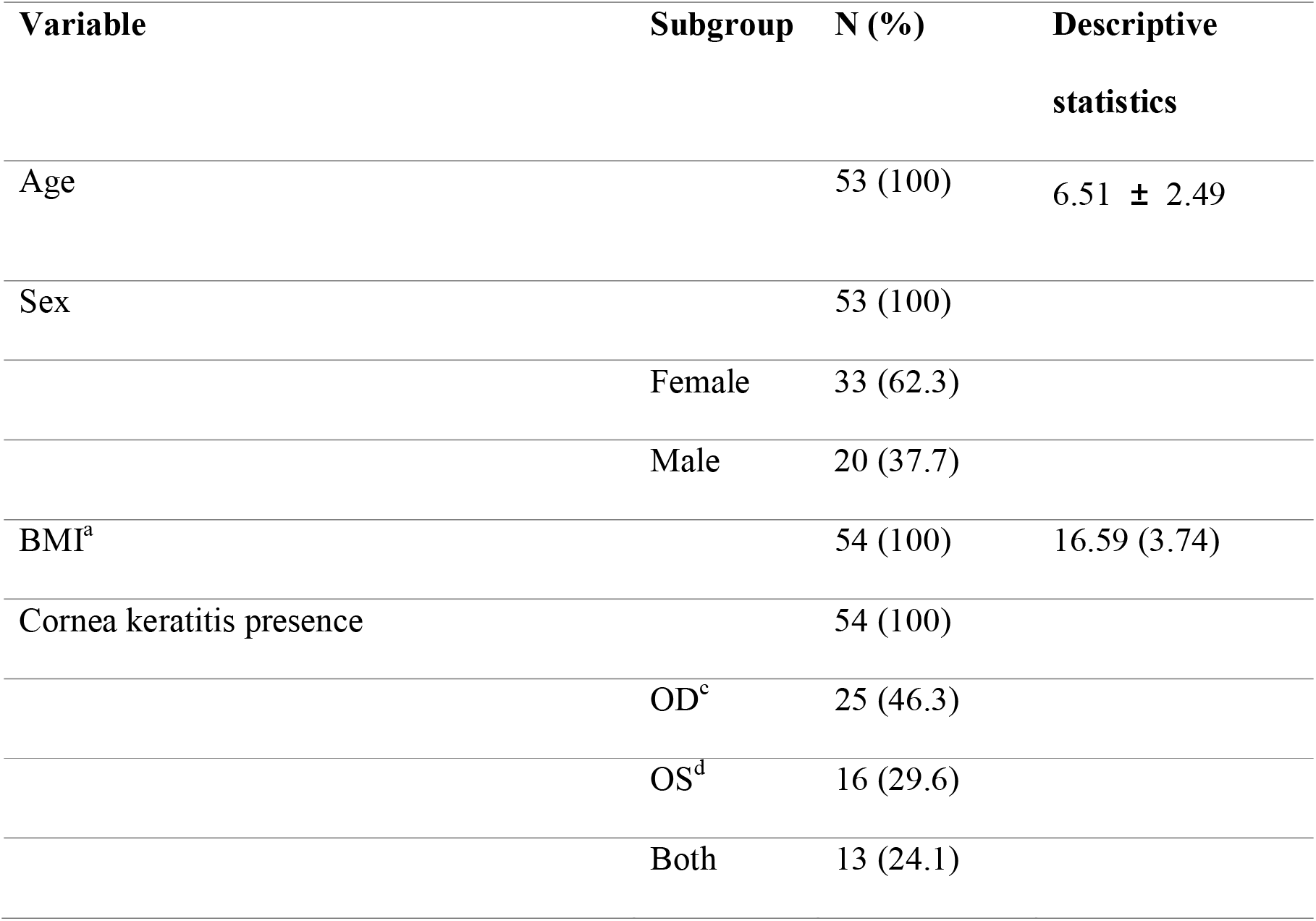
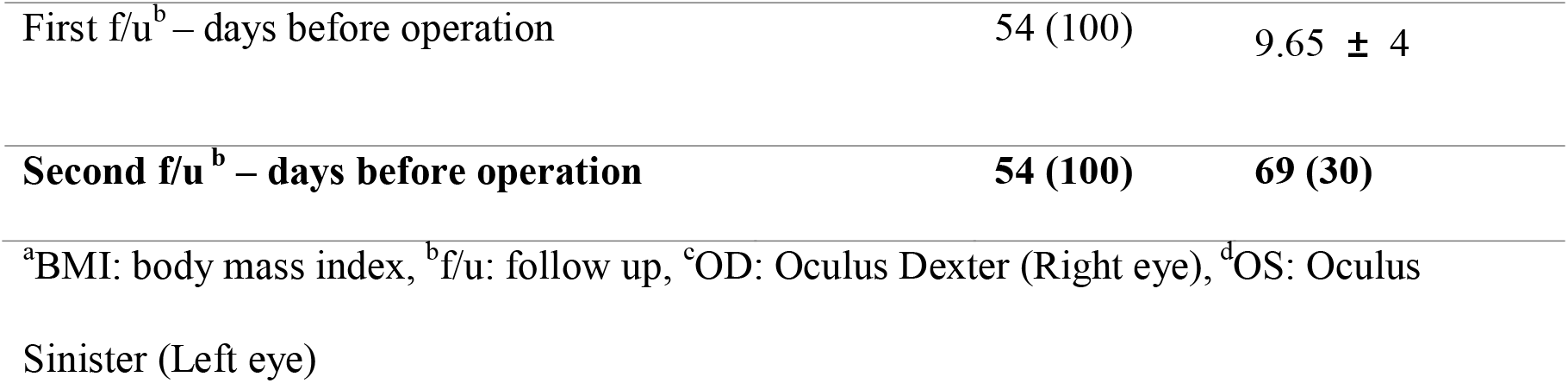
Baseline patient characteristics.

| Variable | Subgroup | N (%) | Descriptive statistics |
| --- | --- | --- | --- |
| Age |  | 53 (100) | 6.51 ± 2.49 |
| Sex |  | 53 (100) |  |
|  | Female | 33 (62.3) |  |
|  | Male | 20 (37.7) |  |
| BMI<sup>a</sup> |  | 54 (100) | 16.59 (3.74) |
| Cornea keratitis presence |  | 54 (100) |  |
|  | OD<sup>c</sup> | 25 (46.3) |  |
|  | OS<sup>d</sup> | 16 (29.6) |  |
|  | Both | 13 (24.1) |  |

|  |  |  |
| --- | --- | --- |
| First f/u<sup>b</sup> – days before operation | 54 (100) | 9.65 ± 4 |
| **Second f/u<sup>b</sup> – days before operation** | **54 (100)** | **69 (30)** |

<sup>a</sup>BMI: body mass index, <sup>b</sup>f/u: follow up, <sup>c</sup>OD: Oculus Dexter (Right eye), <sup>d</sup>OS: Oculus Sinister (Left eye)

Normally distributed variables are expressed as mean ± standard deviation. Non-normally distributed variables are expressed as median (interquartile range). For categorical variables, data are expressed as sample number and percentage, N (%).

Baseline values for LOA and HOA variables are shown in Table 2. In terms of post-surgical changes for LOA variables, flat K was significantly lower at first f/u (*p*=0.023) and 3 months (*p*=0.034) compared to baseline (Supplementary Fig. 1), and mean K values were only significantly different at the second f/u (*p*=0.034; Supplementary Fig. 2). Astigmatism values were significantly higher at second f/u (*p*=0.026) and 1 month (*p*=0.026) compared to baseline (Supplementary Fig. 3), while sphere values were significantly lower at 1 month (*p*=0.041; Supplementary Fig. 4). Axis values were significantly lower at all post-operative time points compared to baseline (*p* ≤ 0.001; Supplementary Fig. 5). No significant changes across time points were observed for steep K (Supplementary Fig. 6), SE (D) (Supplementary Fig. 7), or cylinder values (Supplementary Fig. 8, Table 3).

**Table 2.** Baseline distribution of LOA and HOA variables.

| Type | Variable | Mean ± SD<sup>h</sup> |
| --- | --- | --- |
| <sup>a</sup>LOA | Flat K<sup>b</sup> (D<sup>g</sup>) | 41.82 ± 1.49 |
|  | Steep K (D) | 43.41 ± 1.45 |
|  | Mean K (D) | 42.61 ± 1.30 |
|  | SE<sup>c</sup> (D) | -0.25 (1.76) |
|  | Astigmatism (D) | 1.24 (1.14) |
|  | Cylinder (D) | -0.75 (1.25) |
|  | Sphere (D) | 0.25 (1.75) |
|  | Axis (°) | 180 (90) |
| HOA<sup>d</sup> | Total RMS<sup>e</sup> (D) | 1.23 ± 0.53 |
|  | Total RMS<sup>e</sup> (μm) | 1.60 ± 0.69 |
|  | Defocus (D) | 0.09 ± 0.29 |
|  | 2<sup>nd</sup> Astigmatism (D) | 1.01 ± 0.61 |
|  | Coma (D) | 0.33 ± 0.15 |
|  | Trefoil (D) | 0.17 (0.16) |
|  | **SA<sup>f</sup> (D)** | **-0.11 ± 0.06** |

<sup>a</sup>LOA: low order aberration, <sup>b</sup>K: keratometry, <sup>c</sup>SE: spherical equivalent, <sup>d</sup>HOA: high order aberration, <sup>e</sup>RMS: root mean square, <sup>f</sup>SA: spherical aberration, <sup>g</sup>D: Diopter, <sup>h</sup>SD: standard deviation. Normally distributed variables are expressed as mean ± SD
Non-normally distributed variables are expressed as median (interquartile range)

**Table 3.**
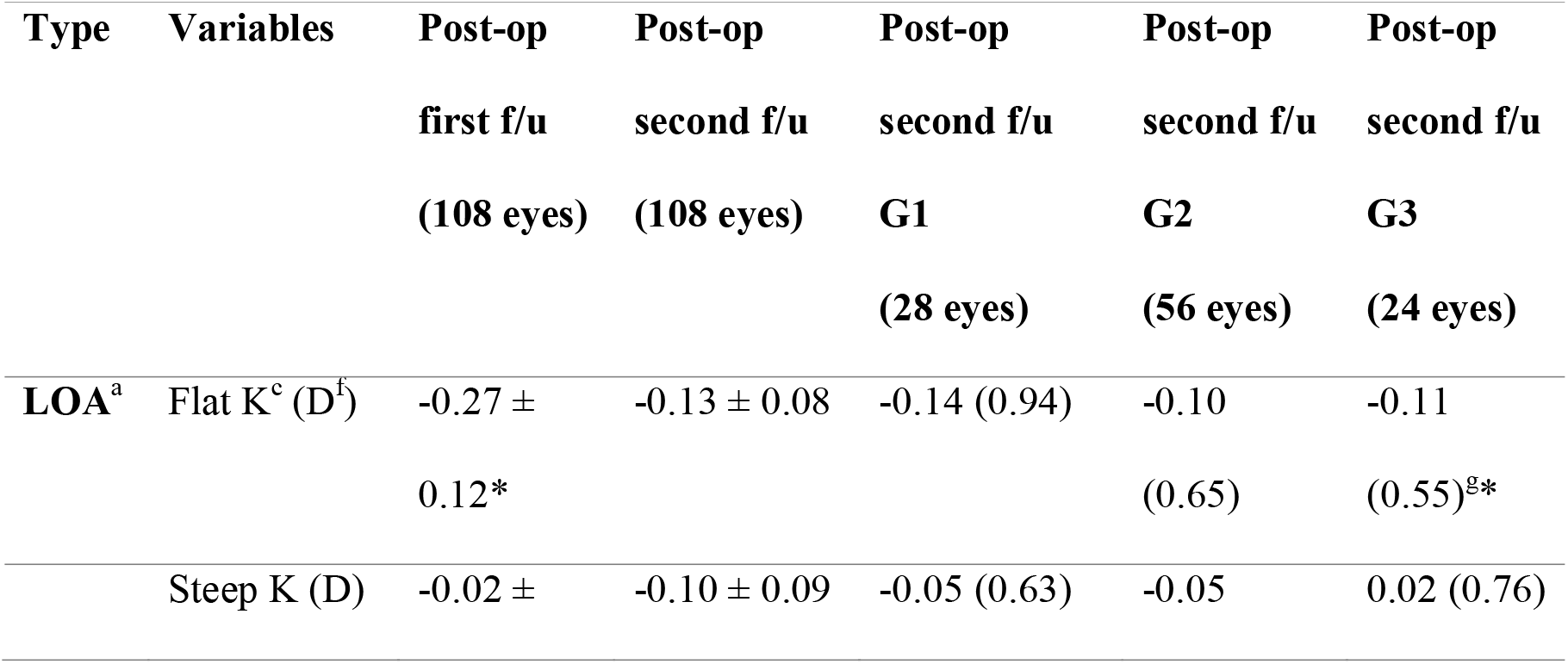
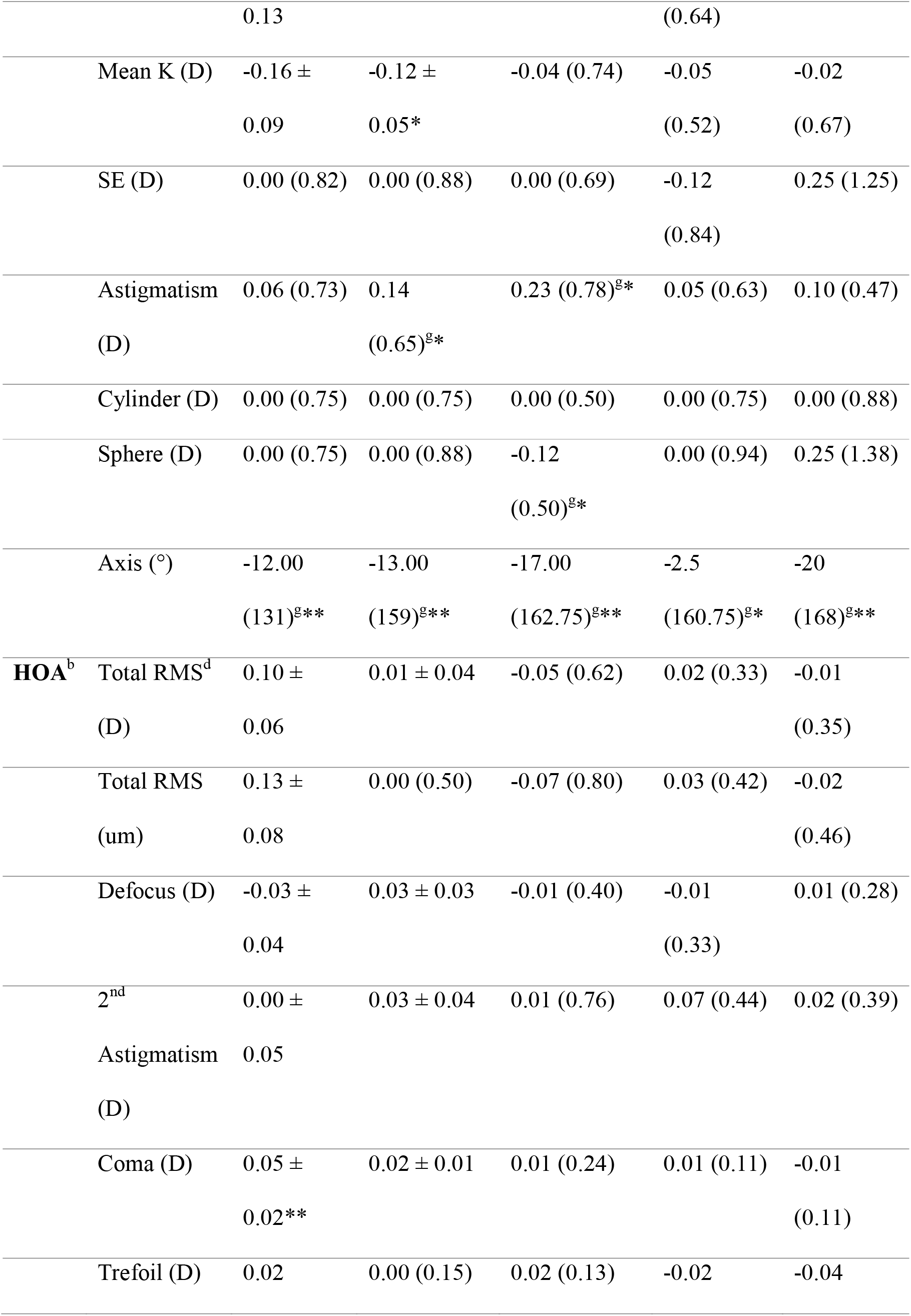
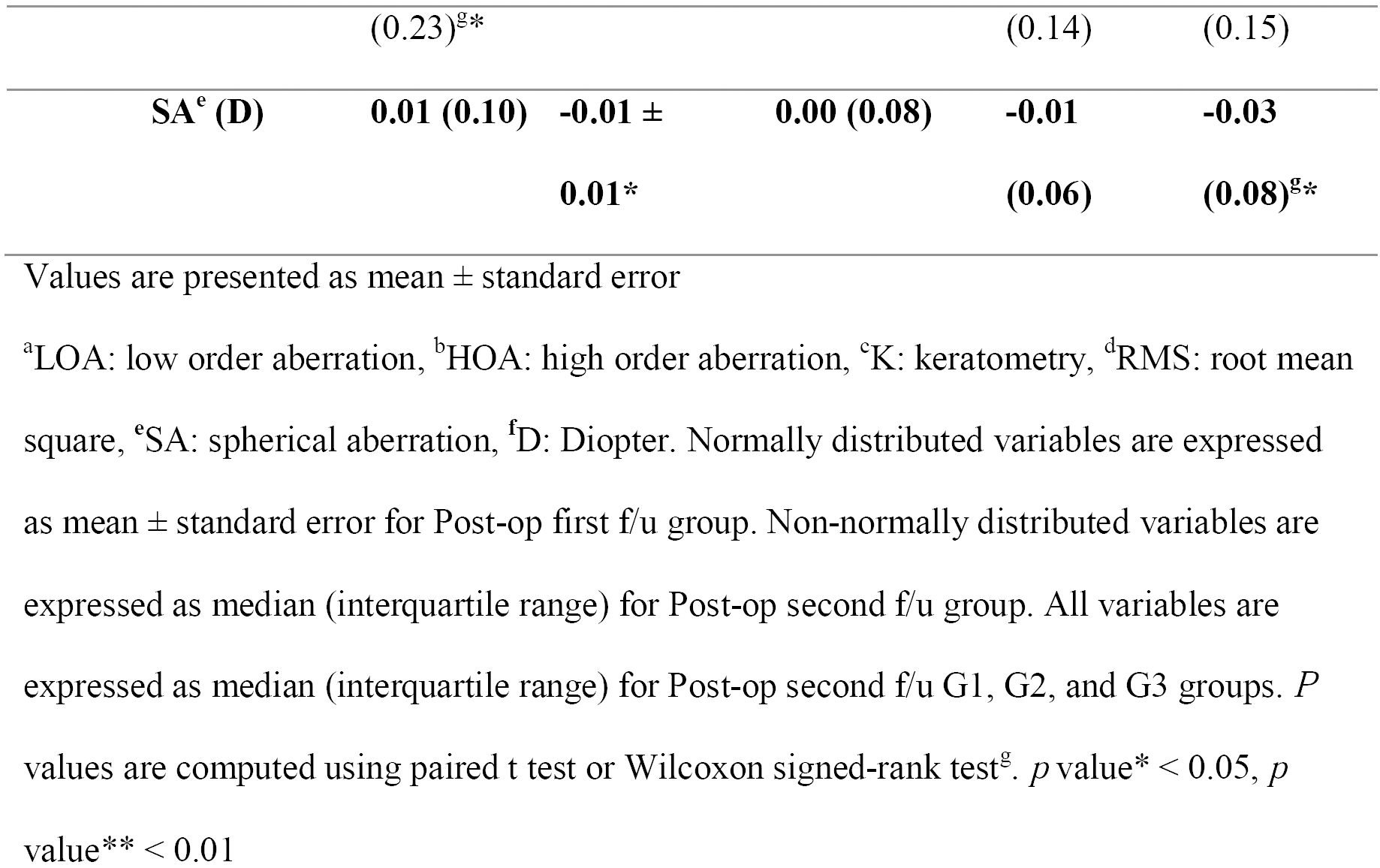
Post-surgical changes in LOA and HOA variables compared to baseline.

In terms of corneal HOAs, both coma (D) (*p*=0.006; Supplementary Fig. 9) and trefoil (D) (*p*=0.037; Supplementary Fig. 10) values were significantly higher at first f/u compared to baseline. No significant differences at any of the post-operative time points, compared to baseline, were observed for SA (Supplementary Fig. 11), total RMS (D) (Supplementary Fig. 12), total RMS (μm) (Supplementary Fig. 13), Defocus (Supplementary Fig. 14), or 2^nd^ Astig values (Supplementary Fig. 15, Table 3).

Univariable linear regression analyses were performed to study the effects of risk factors on postoperative outcomes. At the first and second f/u, sex had an effect on all LOA values except first f/u steep K. Additionally, sex had an effect on all values in G3. Age and BMI affected SE in G1, while sex, age, and BMI affected the first and second f/u times in G2. Sex had an effect on astigmatism values only in G2. The presence of cornea keratitis affected second f/u cylinder values in G3, while sex affected these in G2. Sex, age, and BMI further affected first and second f/u sphere values, while age and BMI affected G1 sphere values, and sex and age affected G2 sphere values. The risk factors assessed did not affect axis values at any time point.

In terms of HOAs, total RMS and RMS were affected by the presence of cornea keratitis at the second f/u only. Age affected defocus values at the second f/u in G2 and G3. However, 2^nd^ Astig was not affected by any risk factors at any time point. Age only affected coma values at the second f/u and in G1, while sex, age, and BMI affected them in G2. Only sex had an effect on trefoil values in G3. Finally, age affected SA values in G2 alone.

Next, we analyzed the significance of time as a fixed effect after correcting for the effects of the risk factors discussed above on the two outcomes (change from baseline values and univariable regression analysis) measured before and after surgery. To do this, a multivariable linear mixed-effect model was employed. At the first f/u, cylinder, coma, trefoil, and SA were significantly increased (*p*=0.039, 0.008, 0.027, and 0.05), while axis and flat K decreased (*p*<0.001, 0.022) from baseline. At the second f/u, cylinder was increased (*p*=0.05), while axis and mean K were significantly decreased (*p*≤0.001 and 0.045) from baseline. In G1, sphere, axis, and flat K decreased from baseline (*p*=0.041, <0.001, and 0.028), while astigmatism increased significantly (*p*=0.028). In G2, axis decreased from baseline (*p*=0.001), while coma increased significantly (*p*=0.04). In G3, axis, flat K, and SA all significantly decreased from baseline (*p*≤0.001, 0.009, and 0.011, respectively).

## DISCUSSION

Given the current deficiencies in the understanding of visual impairments caused by surgery in patients with epiblepharon, this study aimed to investigate post-operative changes in corneal LOAs and HOAs after lower eyelid epiblepharon repair in chlidren. This surgical procedure was demonstrated to cause significant changes in axis, flat K, mean K, SA, coma, and trefoil values.

Many kinds of postoperative visual disturbances due to refractive power changes after ocular surgery have been reported previously. Specifically, severe or irregular astigmatism, changes in the astigmatic axis, myopic or hyperopic shifts, changes in axial length, and positional or dioptric errors in the implanted intraocular lenses have been attributed to visual disturbances and refractive changes in the postoperative period.[21-26] ‘With-the-rule’ astigmatism occurs frequently in epiblepharon patients, as the present paper demonstrates.

The prevalence of ‘with-the-rule’ astigmatism has been reported to range from 60.7% to 90.5% when defined as an axis of astigmatism of 180 ± 15° or 20° minus the cylinder astigmatism axis.[7-9, 27, 28] Here, we report changes from ‘with-the-rule’ to ‘against-the-rule’ astigmatism from 138.26° to 86.05° at 1^st^ f/u and 85.93° at 2^nd^ f/u. (Fig. 5) Furthermore, flat K and mean K decreased after surgery in the present study. Astigmatisms in patients with epiblepharon are thought to result from corneal curvature changes due to the mechanical force caused by abnormal eyelid tension. This tension is caused by redundant horizontal skin folds in the hypertrophied pretarsal orbicularis muscle and squeezing or spasms of the eyelids caused by corneal irritation. On the other hand, Shin et al. found that more severe corneal involvement in epiblepharon was associated with more astigmatism, which persisted after corneal erosions healed.[9] Khwarg et al. further reported that greater corneal injury or a higher number of cilia touching the cornea was correlated with more severe astigmatism. It is hypothesized that long-term corneal epithelial erosion induces cellular apoptosis and cytokine release, resulting in tissue remodeling and keratoconus.[29, 30] Per this model, long-term erosion at the inferior cornea and the mechanical force of the eyelid triggers corneal remodeling and induces astigmatism.

Over the course of the first two post-operative assessments, a trend was observed for the decrease in axis, flat K, and mean K values. This may indicate decreased lower eyelid mechanical force and the presence of cornea keratitis in epiblepharon patients. Vertical pressure by redundant skin folds in the hypertrophied pretarsal orbicularis oculi muscles on the cornea may have resulted in horizontal flattening of the cornea, which could explain the high prevalence of with-the-rule astigmatism in these cases, and flat K change immediately after epiblepharon repair surgery. Conversely, cylinder values were unaffected by epiblepharon surgery. Flat K returned to baseline values in G1 and G2, although a significant drop was observed in G3, and mean K was significantly decreased only at the second f/u. Flat K was more affected by mechanical eyelid forces, while mean K was more affected by corneal erosion.

Correcting for the effects of multiple risk factors (age, sex, BMI, and cornea keratitis presence) on post-surgical outcomes using a multivariable linear mixed-effect model revealed increased cylinder and a significantly decreased flat K at the first f/u. Cylinder and mean K were also significantly different from baseline at the second f/u. This may be because cylinder and keratometry were slightly affected by epiblepharon repair surgery after correcting for LOA risk factors. Given that we used the Galilei G4 Dual Scheimpflug Analyzer to correct for both anterior and posterior astigmatisms, the present study may more accurately reflect pre- to post-epiblepharon surgery topography changes than simulated keratometry approaches that were used previously. Previous studies have reported astigmatic amblyopia in 6-9% of epiblepharon patients.[7, 28] Therefore, in children who have undergone surgical procedures such as epiblepharon repair or strabismus surgery, close monitoring of any refractive changes in the postoperative period is crucial in preventing the development of amblyopia.

HOAs are generally an index of visual quality, including in children.[31] As ocular aberrations increase during development, visual disturbances including distortion and the appearance of glare and halos may occur. However, correction of HOAs improves contrast sensitivity and visual quality. For instance, previous work has found that total HOAs decrease significantly after epiblepharon repair surgery. Critically, the correlation between reduced corneal staining grade and decreased total HOAs may inform the timing of surgery in children without specific visual disturbance and photopobia.[32] However, the present study found that coma and trefoil were significantly increased after epiblepharon repair surgery at the first f/u, and normalized by the second f/u rather than being decreased, supporting previous results.[32] In fact, we found that only SA decreased at the second f/u (especially in G3). Notably, HOAs cannot be corrected by ordinary glasses or contact lenses. Therefore, this indicates that correcting a lens with an astigmatism about 10 days after surgery may not be optimal. About 2 months after epiblepharon surgery may be the optimal time to use correcting lenses.

When accounting for multiple risk factors using a linear mixed-effect model, changes induced by surgery, coma, trefoil, and SA were increased at the first f/u. However, these measures returned to baseline by the second f/u. In G2, only coma increased relative to baseline, while in G3, only SA decreased relative to baseline. This means that coma and trefoil might be affected by the immediate mechanical force changes after surgery, while SA was affected by cornea healing post-operation, especially more than 75 days after surgery.

Recent studies have found that HOAs were significantly more common in patients with dry eye syndrome than in those with healthy eyes,[33,34] and total ocular HOAs were significantly more common in those with dry eyes and central superficial punctuate keratitis than in those with dry eyes alone.[35] Ocular dryness and eye conditions may have a large effect on the outcomes reported here, as indicated by prior studies on HOAs.[33-35] In this study, we found that HOAs generally increased after epiblepharon repair and then almost immediately returned to baseline values. However, these post-surgical results likely vary depending on eye management with hyaluronic acid and other variables such as eye dryness. We were limited by an inability to assess tear film break up time (TBUT) due to the retrospective nature of the present study. It is suggested that future studies consider controlling for this variable when assessing outcomes following epiblepharon surgery. An additional limitation of the present study was the small sample size, which makes it difficult to generalize our results to a broader range of clinical populations. Future prospective studies with larger numbers of patients for postoperative eyelid changes following epiblepharon surgery are needed. Future studies should also investigate the factors that may lead to high levels of aberrations, such as dry eye syndrome and cornea erosion state by using non-clinical (e.g., animal) models.

In conclusion, epiblepharon repair surgery may result in a shift from ‘with-the-rule’ axis changes to ‘against-the-rule’ axis changes. Additionally, flat K, coma, and trefoil were affected by mechanical force changes immediately following surgery. Mean K and SA may also change with cornea state changes in the postoperative period, including corneal erosion healing.

## Data Availability

All data relevant to the study are included in the article or uploaded as supplementary information.

## Acknowledgments

We would like to thank Editage (www.editage.co.kr) for the English language editing

